# Clinical predictors of donor antibody titer and correlation with recipient antibody response in a COVID-19 convalescent plasma clinical trial

**DOI:** 10.1101/2020.06.21.20132944

**Authors:** Maria Lucia L. Madariaga, Jenna J. Guthmiller, Stephen Schrantz, Maud O. Jansen, Chancey Christensen, Madan Kumar, Micah Prochaska, Geoffrey Wool, Amy Durkin-Celauro, Won Hee Oh, Laura Trockman, Janani Vigneswaran, Robert Keskey, Dustin G. Shaw, Haley Dugan, Nai-Ying Zheng, Mari Cobb, Henry Utset, Jiaolong Wang, Olivia Stovicek, Cindy Bethel, Scott Matushek, Mihai Giurcanu, Kathleen G. Beavis, Diego di Sabato, David Meltzer, Mark K. Ferguson, John P. Kress, Kumaran Shanmugarajah, Jeffrey B. Matthews, John F. Fung, Patrick C. Wilson, John C. Alverdy, Jessica S. Donington

## Abstract

**Background:** Convalescent plasma therapy for COVID-19 relies on the transfer of anti-viral antibody from donors to recipients via plasma transfusion. The relationship between clinical characteristics and antibody response to COVID-19 is not well defined. We investigated predictors of convalescent antibody production and quantified recipient antibody response in a convalescent plasma therapy clinical trial.

**Methods:** Multivariable analysis of clinical and serological parameters in 103 confirmed COVID-19 convalescent plasma donors 28 days or more following symptom resolution was performed. Mixed effects regression models with piecewise linear trends were used to characterize serial antibody responses in 10 convalescent plasma recipients with severe COVID-19.

**Findings:** Mean symptom duration of plasma donors was 11.9±5.9 days and 7.8% (8/103) had been hospitalized. Antibody titers ranged from 0 to 1:3,892 (anti-receptor binding domain (RBD)) and 0 to 1:3,289 (anti-spike). Multivariable analysis demonstrated that higher anti-RBD and anti-spike titer were associated with increased age, hospitalization for COVID-19, fever, and absence of myalgia (all p<0.05). Fatigue was significantly associated with anti-RBD (p=0.03) but not anti-spike antibody titer (p=0.11). In pairwise comparison among ABO blood types, AB donors had higher anti-RBD titer than O negative donors (p=0.048) and higher anti-spike titer than O negative (p=0.015) or O positive (p=0.037) donors. Eight of the ten recipients were discharged, one remains on ECMO and one died on ECMO. No toxicity was associated with plasma transfusion. After excluding two ECMO patients and adjusting for donor antibody titer, recipient anti-RBD antibody titer increased on average 31% per day during the first three days post-transfusion (p=0.01) and anti-spike antibody titer by 40.3% (p=0.02).

**Interpretation:** Advanced age, fever, absence of myalgia, fatigue, blood type and hospitalization were associated with higher convalescent antibody titer to COVID-19. Despite variability in donor titer, 80% of convalescent plasma recipients showed significant increase in antibody levels post-transfusion. A more complete understanding of the dose-response effect of plasma transfusion among COVID-19 patients is needed to determine the clinical efficacy of this therapy.

**Trial Registration:** NCT04340050

**Funding:** Department of Surgery University of Chicago, National Institute of Allergy and Infectious Diseases (NIAID) Collaborative Influenza Vaccine Innovation Centers (CIVIC) contract 75N93019C00051

## INTRODUCTION

Convalescent plasma therapy has historically been used as a treatment during epidemics (1). In this therapy, neutralizing anti-viral antibodies, as well as non-neutralizing antibodies and other immunomodulators, are transferred via plasma transfusion from those who have recovered from disease to those currently infected (2-4). For patients with severe COVID-19, convalescent plasma therapy has safely led to improvement in clinical and radiographic parameters (5-10). Once adequate numbers of people convalesced and supply chain logistics were established, providing plasma therapy to a large number of patients has proven feasible (11).

Efficacy of convalescent plasma therapy relies on a robust antibody response in convalescent plasma donors. Measurements of antibody response among patients with COVID-19 demonstrate that the majority develop IgM and IgG within 2 weeks of symptom onset, with specificity towards receptor binding domain (RBD) and spike protein viral epitopes correlating with virus neutralization (12-14). Strikingly, a small proportion of recovered COVID-19 patients show no detectable antibodies to these epitopes (12, 15).

The relationship between host characteristics, disease course and variability in antibody response to COVID-19 is poorly understood. The aim of this study was to establish a translational convalescent plasma program to investigate the relationship between clinical and serological parameters in convalescent plasma donors and define the antibody response of convalescent plasma recipients.

## METHODS

### Study design

This was a prospective open label clinical study to assess the feasibility, safety and immunological impact of delivering anti-SARS-CoV-2 convalescent plasma to hospitalized patients aged 18 years or older with severe or life-threatening COVID-19 disease within 21 days from the onset of their illness. This study was conducted at University of Chicago Medicine (UCM) from April 10, 2020 to May 17, 2020. The final date of follow-up was May 25, 2020.

### Recruitment Team

We used existing hospital infrastructure and personnel to build the convalescent plasma program at a time when state-wide shelter-in-place orders were active, elective procedures were not being performed, and non-COVID-19-related research activities were halted. The donor enrollment team consisted of two surgeons, two surgical residents, and three physician assistants. A dedicated study coordinator was present at the UCM Blood Donation Center to facilitate whole blood donation and collect research samples. Recipients were selected during daily videoconference with infectious disease. One surgeon visited the hospital COVID-19 unit daily to obtain consent and research samples.

### Convalescent Plasma Donors

Plasma donors were age 18 or older, able to donate blood per standard UCM Blood Donation Center guidelines, had a documented COVID-19 polymerase chain reaction (PCR) positive test, and complete resolution of symptoms at least 28 days prior to donation. Recruitment occurred via social media, news outlets, word-of-mouth and announcements in university and community bulletins. The UCM infectious disease team provided an institutional list of patients with a positive PCR test for COVID-19, and their physicians were emailed to request permission to contact the patient for donor participation. Interested plasma donors were directed to fill out a short screening survey online. Potential donors meeting study criteria were screened for eligibility, reported symptoms and comorbidities, consented, and were scheduled for donation at the UCM Blood Donation Center in a single telephone encounter. After meeting the UCM Blood Donation Center eligibility, whole blood was collected and processed according to standard UCM Blood Donation Center procedures. Standard whole blood donation was used for plasma collection because it fit into preexisting UCM Blood Bank infrastructure and workflow therefore facilitating rapid deployment of a collection process, and allowed red blood cell and unused plasma units to be used in the regular Blood Bank inventory. During blood donation, a single research sample was collected at the same time as blood samples for standard immunohematology testing and infectious disease screening. Leukocyte filters used in separation of constituent blood parts were also collected for research.

### Convalescent Plasma Recipients

Eligibility for convalescent plasma recipients included: age 18 or older, laboratory-confirmed COVID-19, within 21 days from the start of illness and severe or life-threatening COVID-19 as defined by the United States Food and Drug Administration (FDA) (16). Severe COVID-19 was defined as dyspnea, respiratory frequency ≥ 30/min, blood oxygen saturation ≤ 93%, partial pressure of arterial oxygen to fraction of inspired oxygen ratio < 300, and/or lung infiltrates > 50% within 24 to 48 hours. Life-threatening COVID-19 was defined as respiratory failure, septic shock, and/or multiple organ dysfunction or failure. Patients who were pregnant, received pooled immunoglobulin in the past 30 days or had a history of transfusion reaction were excluded from this study. Recipients had routine pre-transfusion testing, in keeping with institution policies.

### Convalescent Plasma Transfusion

On the day of enrollment, an emergency investigational new drug (eIND) application was filed and approved for each recipient by the FDA (16). Subsequently, one ABO-compatible unit of convalescent plasma (∼300 mL) was transfused over 4 hours. Repeat administration of convalescent plasma occurred in one recipient (R7). Blood samples and nasopharyngeal swabs were obtained at day 0, 1, 3, 7, 14 post transfusion.

### Outcomes

The primary outcome was feasibility as defined by the collection of convalescent plasma and its administration into hospitalized patients. Secondary outcomes included type and duration of respiratory support, cardiac arrest, transfer to intensive care unit (ICU), length of stay, mortality, complications of plasma administration, process outcomes, and antibody titer of plasma donors and recipients.

Antibody Test and Real-Time Polymerase Chain Reaction (RT-PCR) Detection of SARS-CoV-2 Levels of anti-RBD and anti-spike antibodies were measured by enzyme-linked immunosorbent assay (ELISA) in blood samples at time of donation and plasma recipients, as previously described (17). Nasopharyngeal specimens were obtained by flocked swabs in plasma recipients and analyzed by RT-PCR to detect SARS-CoV-2 RNA.

### Statistics

Study data were collected and managed using REDCap electronic data capture tools hosted at UCM (18, 19). Donor patient characteristics were compared using the chi-squared test for categorical variables and the two-sample t test for continuous variables. Univariate regression analysis for antibody titer (anti-RBD and anti-spike) was conducted against age, sex, body mass index (BMI), previous pregnancy, previous blood donation, blood type, symptoms (fever, cough, sore throat, dyspnea, abdominal pain, aguesia, anosmia, fatigue, myalgia, headache), co-morbidities (respiratory, cardiovascular, renal, diabetes, autoimmune disease, cancer, liver disease), smoking history, travel in the past 3 months to the United States, Asia or Europe, symptom duration, interval from symptoms resolution to plasma donation, and hospitalization. Pairwise comparison using T tests without adjusting for multiple comparisons was used to compare antibody titers among different ABO blood groups.

We conducted multivariable analyses to identify prediction models for anti-RBD and anti-spike antibody titers among convalescent plasma donors. Best subset variable selection method was chosen to identify the subset of predictors that maximizes the adjusted R-squared among all possible models. To compare daily change in recipient antibody response, we fit mixed effects regression models with piecewise linear trend with a change point at 3 days after intervention for log-transformed antibody titers. We considered recipients on extra-corporeal membrane oxygenation (ECMO) (R3 and R6) separately from recipients not on ECMO (R1, 2, 4, 5, 7, 8, 9, 10), because ECMO recipients had different baseline characteristics.

Data analysis was performed using software R, version 3.6.3. Mixed effects regression models were fit using the lmer function of the lme4 package (20). Data analysis was conducted within Rstudio environment, and R markdown files with fully reproducible data analysis can be obtained from the authors upon request.

### Study approval

This study was approved by the Institutional Review Board (IRB20-0523). All participants (plasma donors and plasma recipients) gave written informed consent prior to inclusion in the study. Analysis was performed by MLM and MG. This clinical trial was registered at ClinicalTrials.gov with identifier NCT04340050.

## RESULTS

### Clinical characteristics of convalescent plasma donors

697 potential plasma donors were recruited to our study over 35 days (Table 1). The average age was 43.5 years (range 18 to 87), the majority were female (63.1%), and 37% had never donated blood before. Potential donors with confirmed positive COVID-19 PCR (n=384, 55%) were more likely to be male, have ageusia and anosmia, and lack cough, sore throat and dyspnea compared to the 313 symptomatic patients who had clinical signs of COVID-19 but were never tested (Table 1). Among plasma donors (n=103) who donated as of publication, average symptom duration was 11.9±5.91 days, 9 (8.7%) had respiratory comorbidities such as asthma, chronic obstructive pulmonary disease or obstructive sleep apnea, and 8 (7.8%) had been previously hospitalized for COVID-19 (Table 1). The average interval between symptom start and plasma donation was 45.1±8.02 days.

**Table 1.**
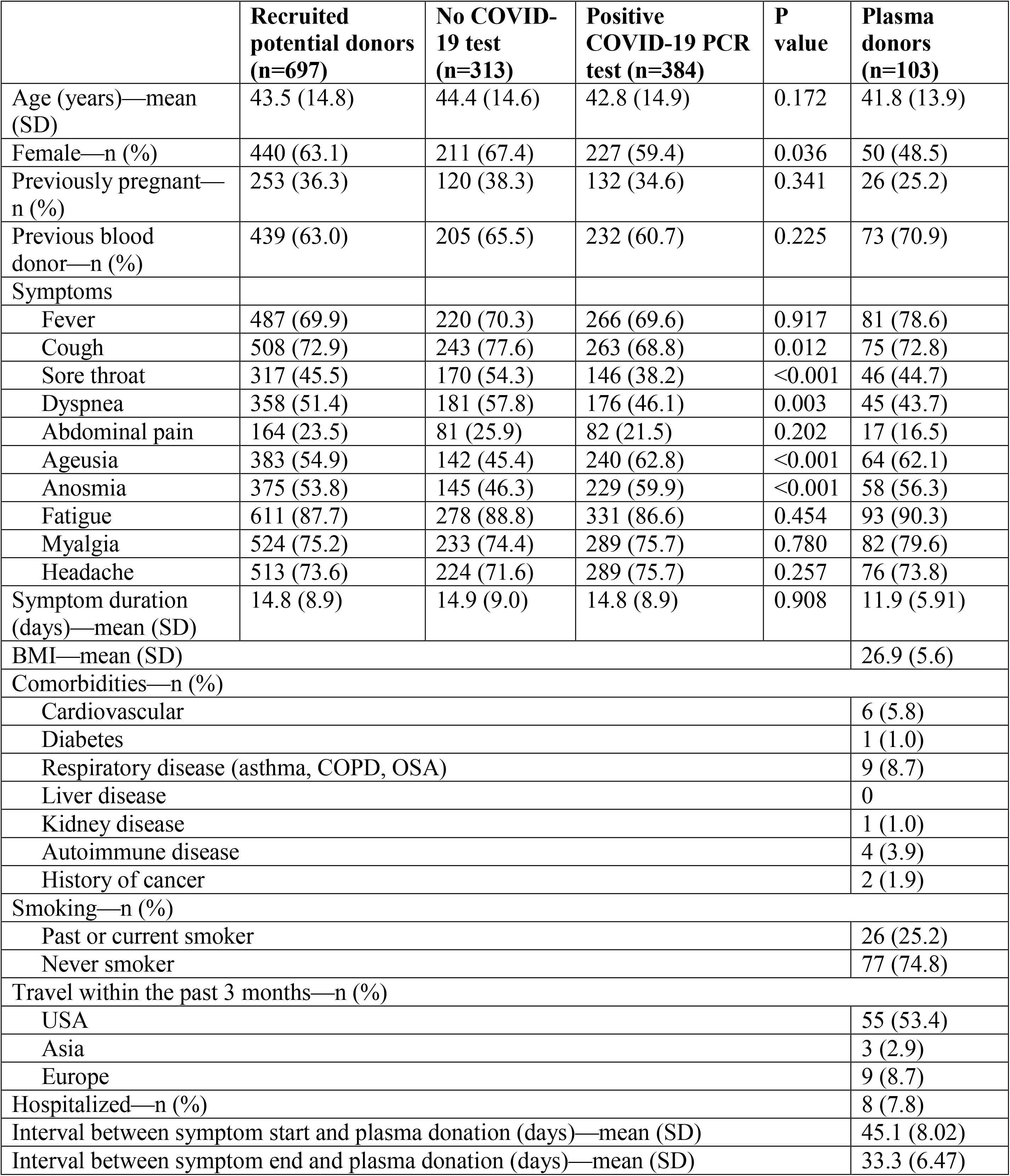
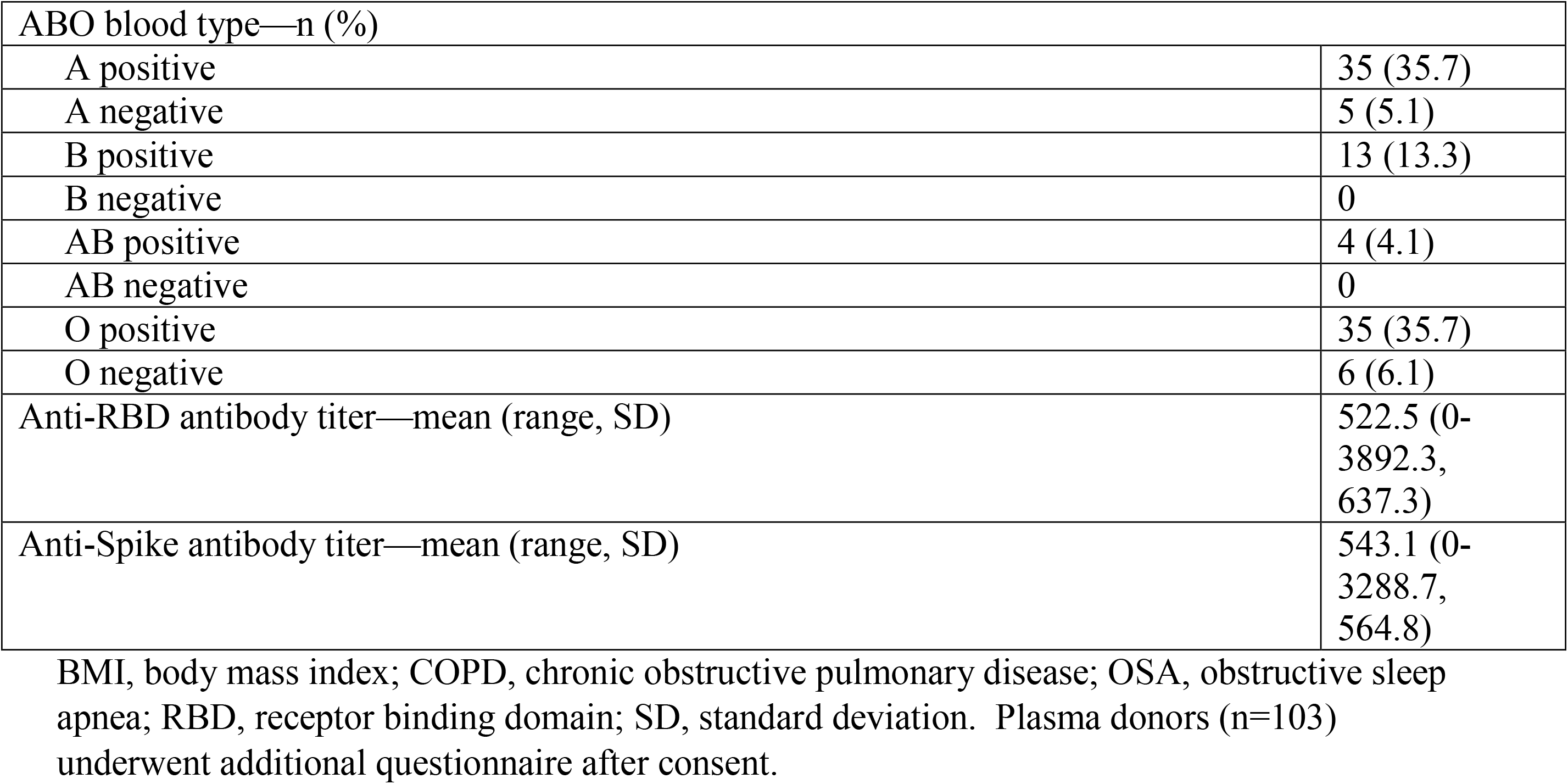
Characteristics of recruited convalescent donors (n=697)

|  | **Recruited potential donors (n=697)** | **No COVID-19 test (n=313)** | **Positive COVID-19 PCR test (n=384)** | **P value** | **Plasma donors (n=103)** |
| --- | --- | --- | --- | --- | --- |
| Age (years)—mean (SD) | 43.5 (14.8) | 44.4 (14.6) | 42.8 (14.9) | 0.172 | 41.8 (13.9) |
| Female—n (%) | 440 (63.1) | 211 (67.4) | 227 (59.4) | 0.036 | 50 (48.5) |
| Previously pregnant—n (%) | 253 (36.3) | 120 (38.3) | 132 (34.6) | 0.341 | 26 (25.2) |
| Previous blood donor—n (%) | 439 (63.0) | 205 (65.5) | 232 (60.7) | 0.225 | 73 (70.9) |
| Symptoms |  |  |  |  |  |
| Fever | 487 (69.9) | 220 (70.3) | 266 (69.6) | 0.917 | 81 (78.6) |
| Cough | 508 (72.9) | 243 (77.6) | 263 (68.8) | 0.012 | 75 (72.8) |
| Sore throat | 317 (45.5) | 170 (54.3) | 146 (38.2) | <0.001 | 46 (44.7) |
| Dyspnea | 358 (51.4) | 181 (57.8) | 176 (46.1) | 0.003 | 45 (43.7) |
| Abdominal pain | 164 (23.5) | 81 (25.9) | 82 (21.5) | 0.202 | 17 (16.5) |
| Ageusia | 383 (54.9) | 142 (45.4) | 240 (62.8) | <0.001 | 64 (62.1) |
| Anosmia | 375 (53.8) | 145 (46.3) | 229 (59.9) | <0.001 | 58 (56.3) |
| Fatigue | 611 (87.7) | 278 (88.8) | 331 (86.6) | 0.454 | 93 (90.3) |
| Myalgia | 524 (75.2) | 233 (74.4) | 289 (75.7) | 0.780 | 82 (79.6) |
| Headache | 513 (73.6) | 224 (71.6) | 289 (75.7) | 0.257 | 76 (73.8) |
| Symptom duration (days)—mean (SD) | 14.8 (8.9) | 14.9 (9.0) | 14.8 (8.9) | 0.908 | 11.9 (5.91) |
| BMI—mean (SD) |  |  |  |  | 26.9 (5.6) |
| Comorbidities—n (%) |  |  |  |  |  |
| Cardiovascular |  |  |  |  | 6 (5.8) |
| Diabetes |  |  |  |  | 1 (1.0) |
| Respiratory disease (asthma, COPD, OSA) |  |  |  |  | 9 (8.7) |
| Liver disease |  |  |  |  | 0 |
| Kidney disease |  |  |  |  | 1 (1.0) |
| Autoimmune disease |  |  |  |  | 4 (3.9) |
| History of cancer |  |  |  |  | 2 (1.9) |
| Smoking—n (%) |  |  |  |  |  |
| Past or current smoker |  |  |  |  | 26 (25.2) |
| Never smoker |  |  |  |  | 77 (74.8) |
| Travel within the past 3 months—n (%) |  |  |  |  |  |
| USA |  |  |  |  | 55 (53.4) |
| Asia |  |  |  |  | 3 (2.9) |
| Europe |  |  |  |  | 9 (8.7) |
| Hospitalized—n (%) |  |  |  |  | 8 (7.8) |
| Interval between symptom start and plasma donation (days)—mean (SD) |  |  |  |  | 45.1 (8.02) |
| Interval between symptom end and plasma donation (days)—mean (SD) |  |  |  |  | 33.3 (6.47) |

|  |  |
| --- | --- |
| ABO blood type—n (%) |  |
| A positive | 35 (35.7) |
| A negative | 5 (5.1) |
| B positive | 13 (13.3) |
| B negative | 0 |
| AB positive | 4 (4.1) |
| AB negative | 0 |
| O positive | 35 (35.7) |
| O negative | 6 (6.1) |
| Anti-RBD antibody titer—mean (range, SD) | 522.5 (0-3892.3, 637.3) |
| Anti-Spike antibody titer—mean (range, SD) | 543.1 (0-3288.7, 564.8) |

BMI, body mass index; COPD, chronic obstructive pulmonary disease; OSA, obstructive sleep apnea; RBD, receptor binding domain; SD, standard deviation. Plasma donors (n=103) underwent additional questionnaire after consent.

### Predictors of donor anti-RBD and anti-spike antibody titer

Donor antibody titers measured on day of plasma donation ranged from 0 to 1:3,892 (anti-RBD) and from 0 to 1:3,288.7 (anti-spike) (Table 1). In univariable regression analysis, higher average anti-RBD and anti-spike antibody titers were associated with plasma donors who were older, male, had higher BMI, had fever and had been hospitalized (p<0.05, Supplemental Table 1). In a pairwise comparison among ABO groups without adjusting for multiple comparisons, AB donors had higher anti-RBD titer than O negative donors (p=0.048) and higher anti-spike titer than O negative (p=0.015) or O positive (p=0.037) donors.

To determine predictors of anti-RBD and anti-spike antibody titer, we performed best subset multivariable analysis including age, sex, blood type, history of previous blood donation, fever, cough, fatigue, myalgia, symptom duration, hospitalization and travel in the United States within the past 3 months. Significant predictors of anti-RBD antibody titer were age (p=0.02), fever (p<0.01), previous hospitalization (p<0.01), lack of myalgia (p=0.01), and fatigue (p=0.03) (R-squared=0.40, adjusted R-squared=0.32, Table 2). Significant predictors of anti-spike antibody titer were age (p=0.02), fever (p=0.01), previous hospitalization (p=0.01), and absence of myalgia (p<0.01) (R-squared=0.35, adjusted R-squared=0.26, Table 2). O positive blood type was associated with lower anti-RBD (p=0.05) but did not meet significance threshold for anti-spike (p=0.07).

**Table 2.** Best subset multivariable regression model for predictors of anti-RBD antibody titer and anti-spike antibody titer.

|  | Reciprocal anti-RBD antibody titer estimate | P value | Reciprocal anti-spike antibody titer estimate | P value |
| --- | --- | --- | --- | --- |
| Age | 10.2 | 0.02 | 9.80 | 0.02 |
| Male | 190.62 | 0.07 | 138.27 | 0.18 |
| Previous blood donation | -213.48 | 0.07 | 137.26 | 0.22 |
| Fever | 362.98 | 0.01 | 347.52 | 0.01 |
| Cough | 178.41 | 0.16 | 180.25 | 0.15 |
| Fatigue | 424.76 | 0.03 | 300.72 | 0.11 |
| Myalgia | -434.68 | 0.01 | -434.56 | <0.01 |
| Symptom duration | -15.36 | 0.11 | -11.2 | 0.23 |
| Hospitalization | 757.60 | <0.01 | 524.8 | 0.005 |
| Travel in USA within 3 months | -149.72 | 0.16 | -188.38 | 0.07 |
| Blood type A negative | -275.97 | 0.29 | -265.37 | 0.29 |
| Blood type O positive | -218.78 | 0.05 | -198.39 | 0.07 |

RBD, receptor binding domain; USA, United States of America. Anti-RBD, R-squared 0.40, adjusted R-squared 0.32. Anti-spike, R-squared 0.35, adjusted R-squared 0.26.

### Clinical course of 10 convalescent plasma recipients

Ten hospitalized patients with severe or life-threatening COVID-19 received plasma on day 0 (Figure 1, Table 3). Plasma recipients were on average 61.9 years old (range 30 to 86) and 40% female. The average time from start of symptoms to plasma transfusion was 12 days (range 2 to 21) and the average time from hospital admission to plasma transfusion was 6 days (range 2 to 17). At the time of plasma transfusion, two patients were on ECMO, one patient was mechanically ventilated, two patients were on high-flow nasal cannula (HFNC), four patients were on nasal cannula and one patient was on room air. Five patients had received other therapies for COVID-19 before transfusion, including remdesivir, tocilizumab, anakinra and hydroxychloroquine. Two plasma recipients were on chronic immunosuppression after transplantation.

**Table 3.** Clinical characteristics of recipients receiving plasma for this study.

| **Respiratory support prior to plasma transfusion** | **Recipient #** | **Age Decade** | **Gender** | **Symptoms** | **Previous COVID treatment** | **Interval from symptom start to plasma transfusion (days)** | **Interval from hospital admission to plasma transfusion (days)** | **Donor Anti-RBD titer (1/X)** | **Recipient Anti-RBD titer (1/X) on day 0** | **Donor Anti-spike titer (1/X)** | **Recipient Anti-spike titer (1/X) on day 0** | **Disposition** |
| --- | --- | --- | --- | --- | --- | --- | --- | --- | --- | --- | --- | --- |
| **Veno-venous ECMO** | R3 | 50 | F | Fever, cough, dyspnea | Remdesivir, tocilizumab | 21 | 14 | 585.4 | 2732.1 | 580.4 | 7666.7 | Died (day 30) |
|  | R6 | 50 | M | Fever, chills, decreased appetite, dizziness | Remdesivir, tocilizumab | 20 | 17 | 78.7 | 4301.6 | 305.1 | 13833.6 | ICU (day 32) |
| **Mechanical ventilation** | R1 | 50 | M | Fever, cough, nausea | Tocilizumab | 10 | 3 | 80.2 | 0 | 68.9 | 0 | Rehab (day 24) |
| **High-flow nasal cannula** | R9 | 70 | M | Fever, cough | none | 14 | 6 | 1312.8 | 879.7 | 1179.2 | 2118.4 | Rehab (day 26) |
| **Nasal Cannula** | R5 | 60 | F | Altered mental status, dyspnea | none | 9 | 2 | 292.4 | 113.6 | 351.1 | 181.6 | Rehab (day 14) |
|  | R7 | 50 | M | Dyspnea | Tocilizumab, anakinra | 9 | 8 | 2330.2 | 810 | 1648 | 1056.6 | Rehab (day 33) |
|  | R10 | 80 | F | Dyspnea, abdominal pain | none | 6 | 2 | 434.8 | 0 | 756.4 | 0 | Home (day 23) |
|  | R2 | 60 | M | Cough, weakness, hiccups, altered mental status | Hydroxy-chloroquine | 16 | 3 | 72.6 | 0 | 92.7 | 446.2 | Home (day 9) |
|  | R4 | 70 | F | Fever, altered mental status | none | 2 | 2 | 382.0 | 1461.3 | 716.3 | 3154.4 | Home (day 8) |
| **Room air** | R8 | 30 | M | Fever, chills,<br>fatigue,<br>ageusia | none | 13 | 3 | 3892.<br>3 | 71.9 | 2921.<br>1 | 0 | Home (day 6) |

AKA, above the knee amputation; CHF, congestive heart failure; DM, diabetes mellitus; DVT, deep venous thrombosis; ESRD, end-stage renal disease; HTN, hypertension; NAFLD, non-alcoholic fatty liver disease; PE, pulmonary embolism; PVD, peripheral vascular disease.

**Figure 1.**
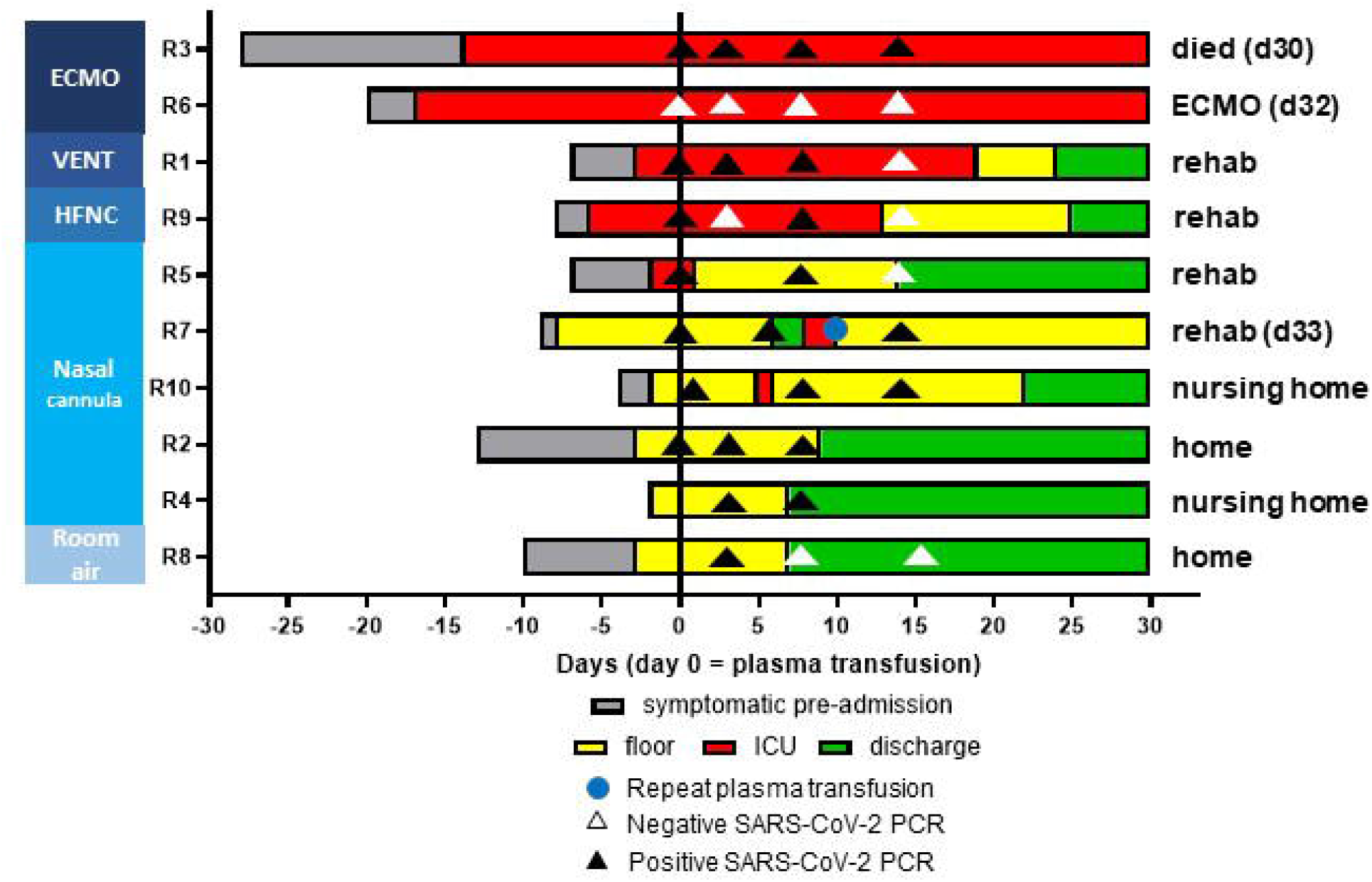
Recipient hospital course. Recipient clinical course before and after plasma transfusion (day 0). Number of days symptomatic prior to admission (gray) and recipient location in the intensive care unit (ICU, red), hospital floor (yellow) and home (green) shown by day of plasma transfusion. Positive SARS-Cov2 NP swab PCR test indicated by black triangle and negative test indicated by white triangle. Repeat plasma dosing indicated by blue circle. Respiratory support at time of plasma transfusion indicated by left column (ECMO, extracorporeal membrane oxygenation; vent, mechanical ventilation; nasal cannula; room air).

Figure 2 shows selected clinical and laboratory parameters of convalescent plasma recipients. Only one recipient (R8) had fever prior to transfusion and this resolved by day 3 post-transfusion. R3 and R6 remained on ECMO throughout the study period. In the remaining 8 recipients, oxygen requirements improved to room air or nasal cannula. The Sequential Organ Failure Assessment (SOFA) score (21) was calculated for recipients on mechanical ventilation or ECMO and showed a general trend towards improvement; notably both ECMO patients were weaned off vasopressor and intra-aortic balloon pump support by 7 days post-transfusion.

**Figure 2.**
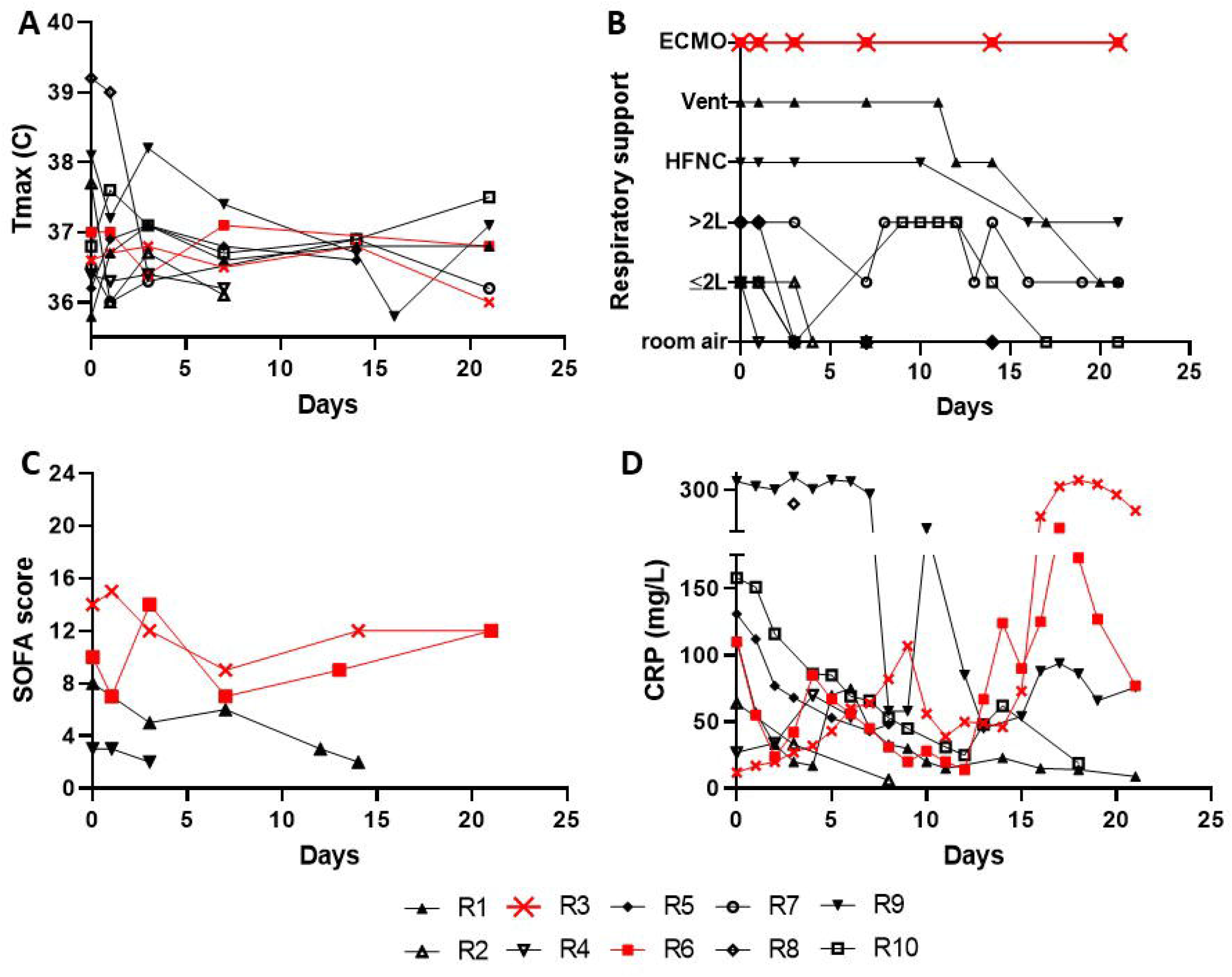
Recipient clinical and laboratory parameters after plasma transfusion. (A) Maximum daily temperature (Tmax, C); (B) Type of respiratory support required (ECMO, extracorporeal membranous oxygenation; Vent, mechanically ventilated; HFNC, high-flow nasal cannula; L, number of liters of oxygen on nasal cannula); (C) Sequential Organ Failure Assessment (SOFA) score for recipients on mechanical ventilation or ECMO; (D) Inflammatory marker C-reactive protein (CRP). Data for patients on ECMO are in red.

Levels of inflammatory marker C-reactive protein (CRP) were variable. CRP decreased in six recipients (R1, R2, R5, R6, R9, R10). SARS-CoV2 NP swab PCR remained positive in 5 patients and turned negative in 4 patients; 1 patient (R6) had been positive for SARS-CoV2 17 days prior to plasma transfusion but was negative for SARS-COV2 on day of transfusion (Figure 1). At last follow-up, 1 patient on ECMO remained in the hospital (R6), 1 patient on ECMO was transitioned to comfort care and died on day 30 after plasma transfusion (R3), 4 patients were discharged to rehabilitation facilities and 4 patients were discharged to their place of residence (Figure 1).

### Post-transfusion relationship between convalescent plasma donor and recipient antibody titer

On day of transfusion, anti-RBD antibody titers were undetectable in 3 recipients (R1, R2, R10) and anti-spike antibody titers were undetectable in 3 recipients (R1, R8, R10) (Table 3 and Figure 3). Both patients on ECMO had very high antibody titer at day 0 which decreased in the days after transfusion (Figure 3). The remaining plasma recipients showed increase in antibody titer within the first three days after transfusion (R1, 2, 4, 5, 7, 8, 9) with the exception of R10 who did not show any antibody titer until day 7 (anti-spike) and day 14 (anti-RBD) after transfusion (Figure 3).

**Figure 3.**
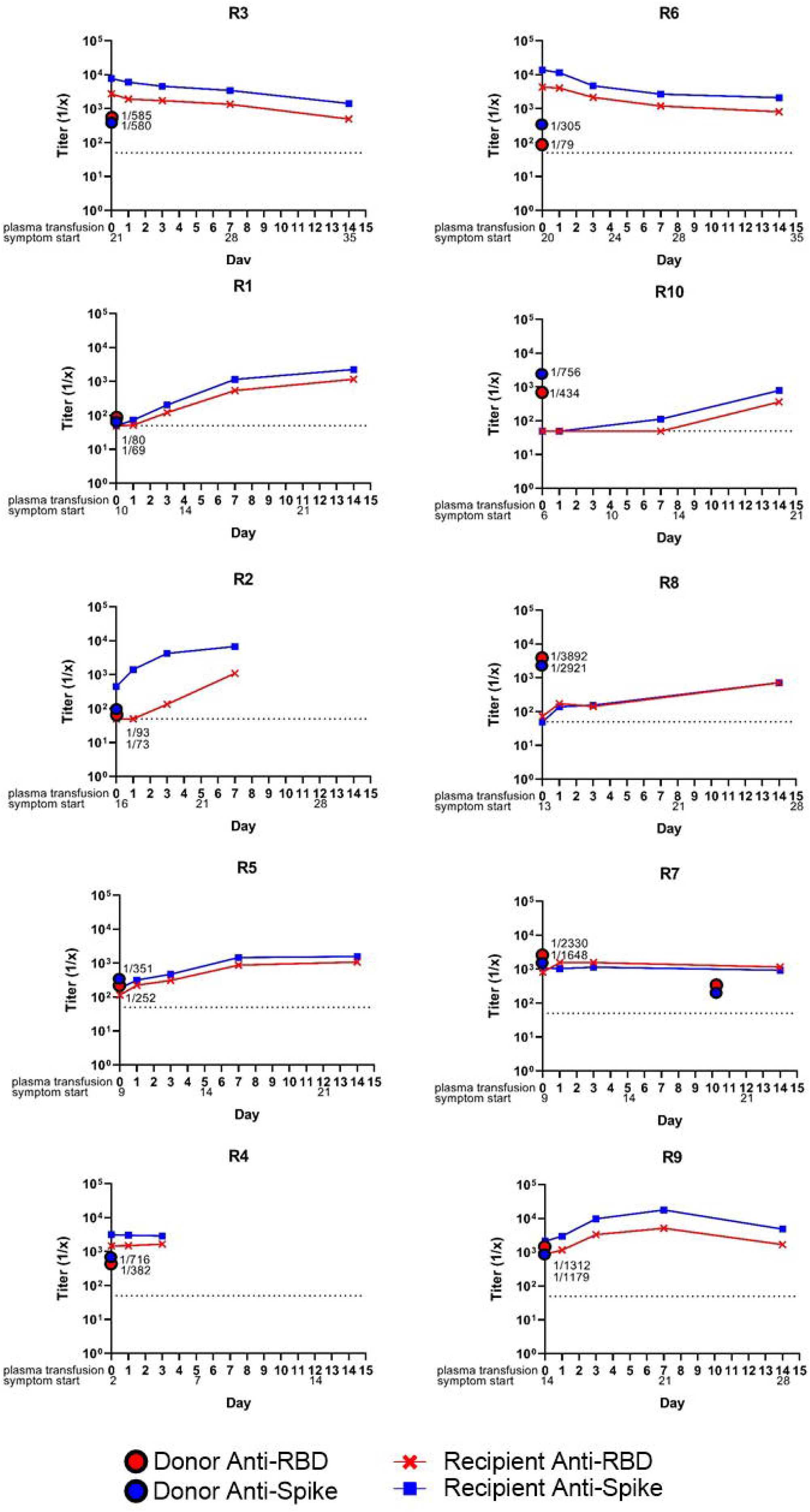
Recipient serology after plasma transfusion. Reciprocal donor plasma anti-RBD (red circle) and anti-spike (blue circle) antibody titer are plotted on the y-axis. Dotted line at 1:50 represents the limits of antibody detection. Reciprocal recipient anti-RBD (red line) and anti-spike (blue line) antibody titer are plotted on day 0 prior to transfusion and on days 1, 3, 7 and 14 post-transfusion.

We performed a mixed effects model for log-transformed reciprocal antibody titer adjusting for donor antibody titer level looking at the first 3 days post-transfusion among the non-ECMO patients. After plasma transfusion, recipient anti-RBD antibody titer increased on average by 31% per day (p=0.01) and recipient anti-spike antibody titer increased on average by 40.3% per day (p=0.01) (Figure 4). Among the two ECMO recipients, recipient antibody response was not significantly changed until three days after plasma transfusion (decreasing by 9.2% per day for anti-RBD titer and 8.2% per day for anti-spike titer, p<0.01) (Figure 4).

**Figure 4.**
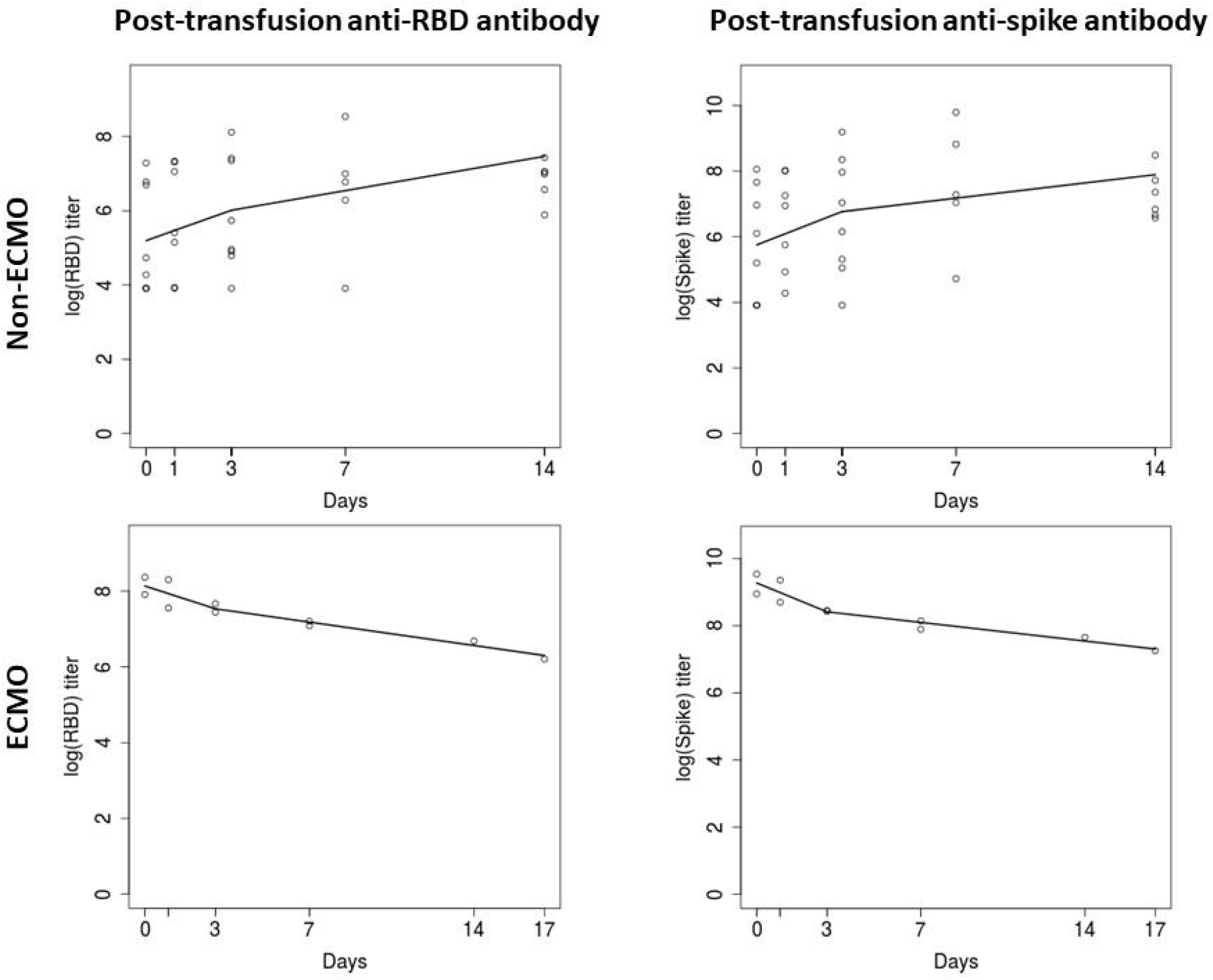
Antibody response curve in plasma recipients. Log-transformed rate of antibody titer change of anti-RBD and anti-spike antibodies in the non-ECMO (n=8) and ECMO (n=2) recipients were fitted in a mixed effects piece-wise linear regression model.

### Safety of convalescent plasma transfusion

We monitored the clinical status of the recipients before, during and immediately after transfusion. No recipients experienced toxicity associated with plasma transfusion. There was no clinical deterioration or worsening of disease status immediately related to plasma transfusion. Convalescent plasma transfusion was safe in high-risk individuals in our study: immunosuppressed patients after stem cell and lung transplants and a patient with end-stage renal disease on dialysis.

## DISCUSSION

We developed a translational convalescent plasma treatment program within the existing hospital infrastructure during the COVID-19 pandemic that provided a new therapeutic option for patients while assessing the antibody profile of both convalescent and hospitalized patient populations. Our multivariable analysis demonstrated that clinical characteristics can predict serological response of antibodies associated with virus neutralization (12). Higher anti-RBD and anti-spike antibody were more likely found in convalescents who were older, hospitalized, had fever, and lacked myalgia. Fatigue also significantly predicted higher anti-RBD but not anti-spike antibody titer.

Variability in convalescent populations and immune response to viral infection may explain why recovery is not always marked by seroconversion (12, 15). Indeed, in our study four plasma donors (as well as four plasma recipients) had undetectable antibody titers.

Disparate plasma donor populations and geography may explain why symptom duration and elapsed time from symptom onset was associated with antibody response in New York City (13) but not among our patients in Chicago. Disparate plasma donor populations and geography may also explain antibody variability. These data highlights that the impact of variability in antibody type and titer on virus-neutralizing activity and long-term immunity is unknown.

Interestingly, we found that antibody titers significantly differed across ABO blood type groups, with O donors (who have natural anti-A and anti-B antibodies) demonstrating lower anti-RBD and anti-spike titers than AB donors. Previous studies showed that O blood type populations are less susceptible to infection with SARS-CoV (22) and SARS-CoV2 (COVID-19) (23). Anti-A antibodies inhibited binding of the SARS-CoV spike protein to angiotensin-converting enzyme 2 receptors *in vitro* (24). Further studies on the relationship between ABO polymorphism and antibody titer may uncover genetic determinants of the host response to COVID-19.

Recipients received plasma with a range of antibody titer from 1:73 to 1:3,892 (anti-RBD) and 1:69 to 1:2,921 (anti-spike). Despite this, 80% of recipients demonstrated a significant increase in anti-spike and anti-RBD antibody titer in the 3 days post transfusion that was independent of donor antibody titer and were discharged after clinical improvement.

Interestingly, recipient antibody titer continued to increase up to 14 days in four recipients (R1, 2, 8, 10); in contrast, the two most severely ill patients on ECMO who had the highest antibody titers (up to 1:13,833 anti-spike antibody in R6) showed a decrease in antibody titer after receiving plasma on day 20-21 of illness.

Importantly we demonstrate the safety of transfusing convalescent plasma in immunosuppressed patients after lung transplantation and stem cell transplantation. None of the plasma recipients in this study deteriorated after convalescent plasma transfusion, consistent with the safety profile of other trials (5-9, 11). Repeat plasma dose in recipient R7 was also well-tolerated. Pre-clinical models of SARS-CoV and clinical experience of other viral illness had raised concern about the potential for non-neutralizing antibody to cause antibody dependent enhancement of disease, which was not seen here despite variable titers of donor antibodies (25-27).

The variability in post-transfusion recipient antibody titer and clinical response seen here and in other studies (5, 6, 28, 29) indicates that the therapeutic activity of convalescent plasma depends on the timing of treatment and composition of convalescent plasma. Indeed, plasma contains more than 1,000 proteins, including albumin, immunoglobulins, complement, and coagulation factors as well as organic compounds such as cytokines (4). Convalescent plasma drawn shortly after natural infection (1, 5-8) may be enriched for populations of protective antibodies not present in plasma derived from long-recovered or rarely-hospitalized donors studied here. Furthermore, immunomodulatory and non-virus neutralizing antibody effects such as stimulation of the host humoral immune response and facilitating viral uptake into cells via Fc-receptors to increase viral antigen presentation to other effector cells may contribute to disease recovery. Taken together, while randomized controlled efficacy trials for convalescent plasma therapy in COVID-19 are currently underway, establishing effective anti-COVID-19 plasma-based therapy will require both an understanding of the precise dose and type of virus-neutralizing antibody and in-depth characterization of plasma donor-recipient pairs.

The availability of a pre-existing hospital-based blood collection facility within our medical center significantly eased the procurement of convalescent plasma and will allow us to assess immunological characteristics of donor-recipient pairs in future studies. Such hospital-based blood collection facilities have been declining in number across the United States for several decades (30). Cultivating region-specific convalescent plasma inventory may potentially facilitate the identification and isolation of antibodies with specific activity against local virus strains and be a useful model for future outbreaks. In addition, convalescent plasma derived from whole blood collection is a rapidly scalable technique that requires basic phlebotomy and blood separation rather than a dedicated apheresis personnel and equipment. Furthermore, a significant proportion (36.3%) of our plasma donors had never donated blood before, indicating that a convalescent plasma donation program can serve as important community outreach during a time when patients avoid hospitals that are perceived as unsafe (31).

In summary, development of a convalescent plasma program is feasible, rapidly deployable and economical when existing resources of equipment, space and personnel are used. Establishing the clinical predictors of high antibody titer and understanding the serological post-transfusion response may guide patient selection and shed light on antibody response to COVID-19. Further work characterizing convalescent plasma donor and recipient pairs is needed to elucidate mechanisms of convalescent plasma therapy and demonstrate optimal viral epitope therapeutic targets.

## Data Availability

All data is available upon request.

## AUTHOR CONTRIBUTIONS

MLM, JSD designed the study, executed the study, acquired and analyzed data, wrote the manuscript.

SS, MK, JPK trial design, recipient selection team, Covid-19 testing.

MP, trial design, donor recruitment and screening.

ADC, WHO, LT, JV, RK donor recruitment and selection.

MOJ study coordinator, blood bank liaison.

CC, GW blood bank, co-ordination, trial design.

CB, SM, KB covid testing.

JJG, DS, HD, NYZ, MC, HU, JW, OS, generation of recombinant proteins, ELISA antibody testing and research sample processing.

DdS, MKF, KS, JBM, JFF, PCW, JCA study design, data analysis, critical review of the manuscript.

## ACKNOWLEDGEMENTS

We thank all the plasma donors for their willingness to help in a time of need and the blood bank staff for their excellent care. We thank Samantha Guerrero, Alyssa Anneken, Bruce Boehrnsen and Rohit Allada for helping us establish the infrastructure for this study. We thank the University of Chicago and Department of Surgery, University of Chicago for providing support for this study. This study was funded by the Department of Surgery, University of Chicago and the National Institute of Allergy and Infectious Diseases (NIAID) Collaborative Influenza Vaccine Innovation Centers (CIVIC) contract 75N93019C00051.

## SUPPLEMENTAL TABLES

**Supplemental Table 1.** Univariate regression analysis for reciprocal anti-RBD and anti-spike antibody titer.

|  | Anti-RBD<br>reciprocal<br>antibody titer<br>estimate | P value | Anti-spike<br>reciprocal<br>antibody titer<br>estimate | P value |
| --- | --- | --- | --- | --- |
| Age | 12.6 | 0.005 | 11.7 | 0.003 |
| BMI | 23.48 | 0.056 | 23.9 | 0.019 |
| Male vs Female | 647.52 vs 390.1 | 0.036 | 660.61 vs 418.64 | 0.026 |
| Fever vs none | 605.2 vs 210.5 | 0.0085 | 614.9 vs 272.2 | 0.01 |
| Hospitalized<br>versus not | 1535 vs 438.9 | <0.001 | 1227.2 vs 486.6 | <0.001 |

BMI, body mass index; RBD, receptor binding domain. Previously pregnant, previous blood donor, cough, sore throat, dyspnea, abdominal pain, ageusia, anosmia, fatigue, myalgia, headache, symptom duration, comorbidities (respiratory, cardiovascular, kidney disease, liver disease, diabetes, autoimmune disease, history of cancer), smoking history, travel history (within the past 3 months to USA, Asia, Europe), interval of symptoms to plasma donation, blood type were not significantly associated with anti-RBD or anti-spike antibody titer.

**Supplemental Table 2.**
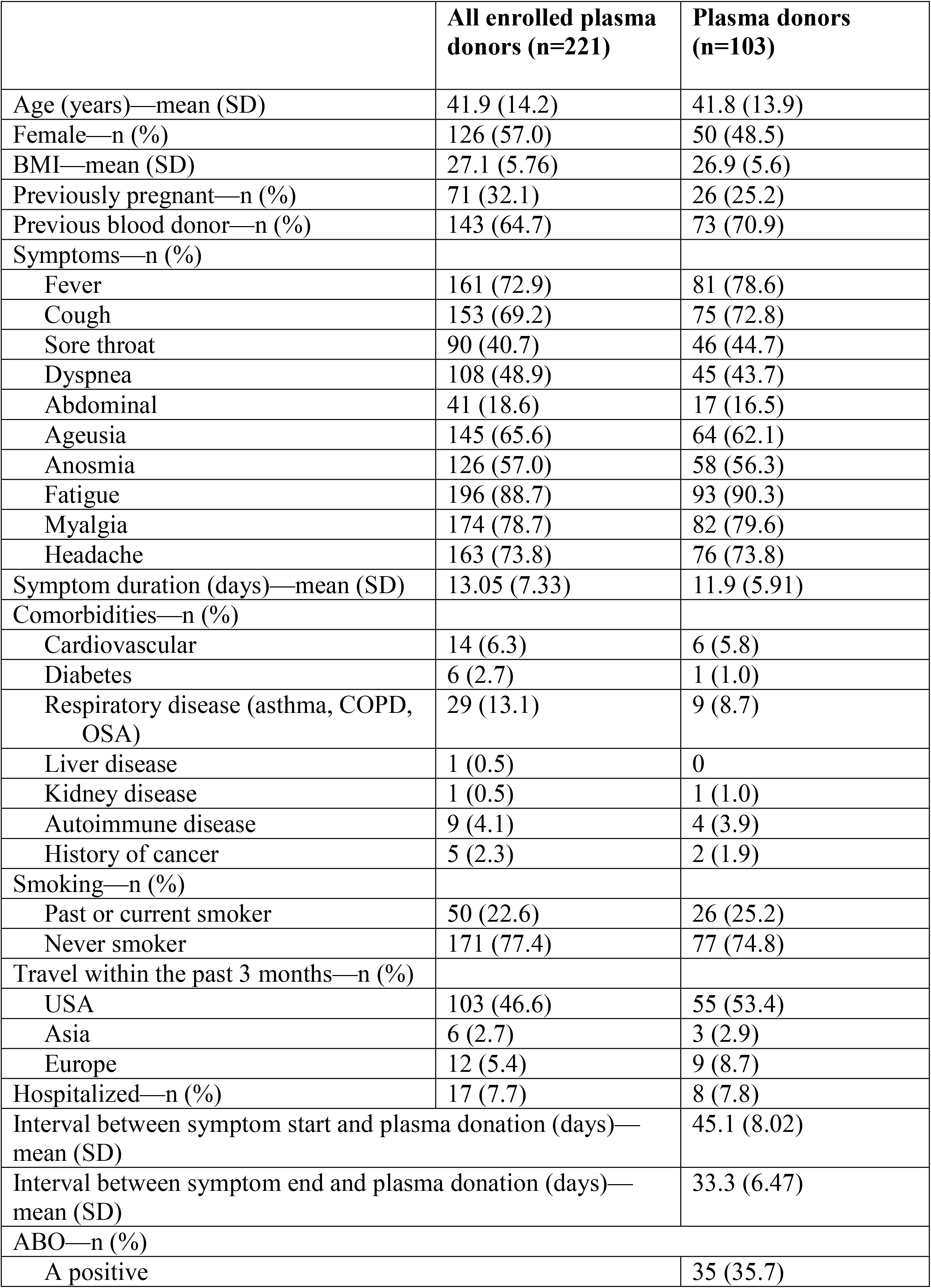
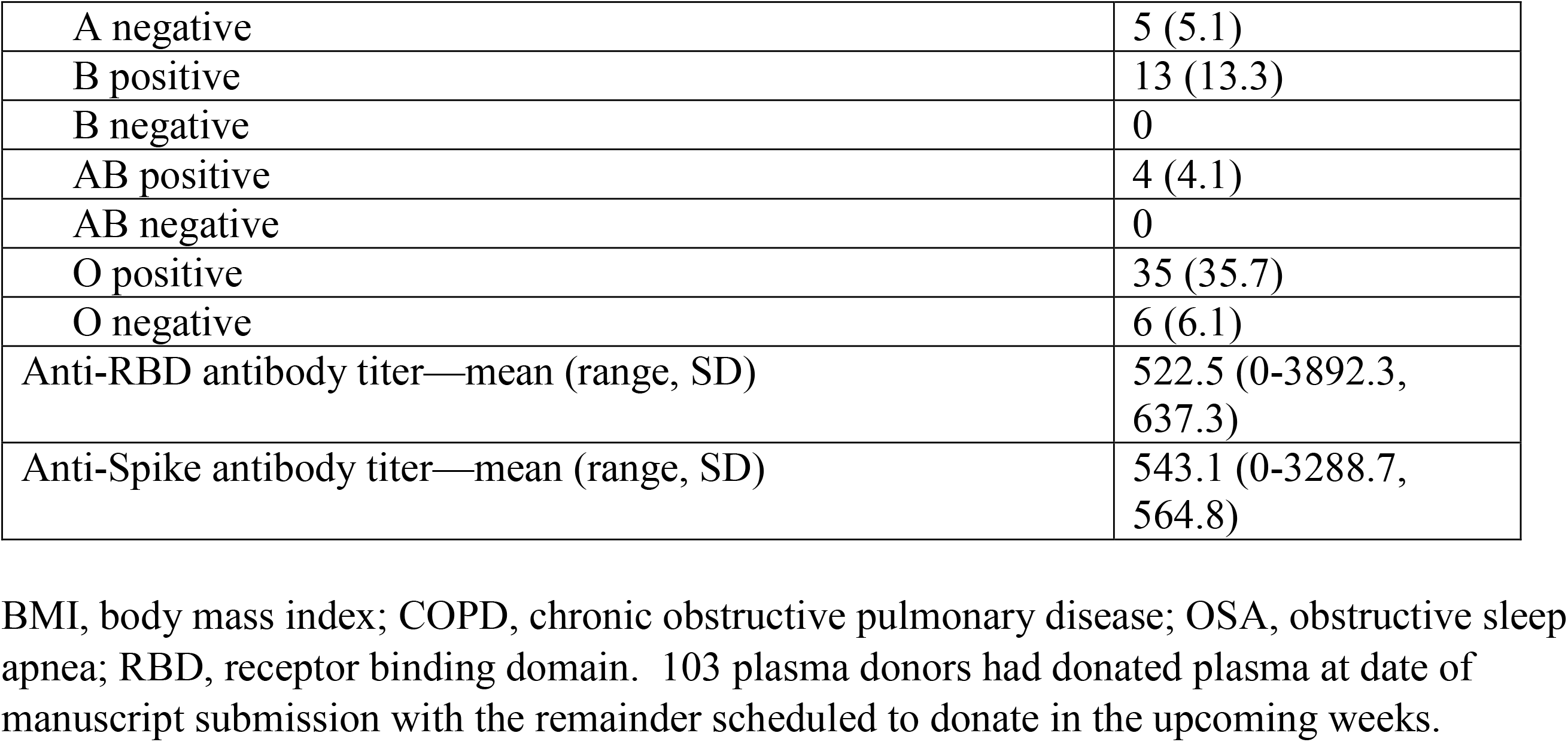
Detailed characteristics of enrolled plasma donors (n=221)

|  | **All enrolled plasma donors (n=221)** | **Plasma donors (n=103)** |
| --- | --- | --- |
| Age (years)—mean (SD) | 41.9 (14.2) | 41.8 (13.9) |
| Female—n (%) | 126 (57.0) | 50 (48.5) |
| BMI—mean (SD) | 27.1 (5.76) | 26.9 (5.6) |
| Previously pregnant—n (%) | 71 (32.1) | 26 (25.2) |
| Previous blood donor—n (%) | 143 (64.7) | 73 (70.9) |
| Symptoms—n (%) |  |  |
| Fever | 161 (72.9) | 81 (78.6) |
| Cough | 153 (69.2) | 75 (72.8) |
| Sore throat | 90 (40.7) | 46 (44.7) |
| Dyspnea | 108 (48.9) | 45 (43.7) |
| Abdominal | 41 (18.6) | 17 (16.5) |
| Ageusia | 145 (65.6) | 64 (62.1) |
| Anosmia | 126 (57.0) | 58 (56.3) |
| Fatigue | 196 (88.7) | 93 (90.3) |
| Myalgia | 174 (78.7) | 82 (79.6) |
| Headache | 163 (73.8) | 76 (73.8) |
| Symptom duration (days)—mean (SD) | 13.05 (7.33) | 11.9 (5.91) |
| Comorbidities—n (%) |  |  |
| Cardiovascular | 14 (6.3) | 6 (5.8) |
| Diabetes | 6 (2.7) | 1 (1.0) |
| Respiratory disease (asthma, COPD, OSA) | 29 (13.1) | 9 (8.7) |
| Liver disease | 1 (0.5) | 0 |
| Kidney disease | 1 (0.5) | 1 (1.0) |
| Autoimmune disease | 9 (4.1) | 4 (3.9) |
| History of cancer | 5 (2.3) | 2 (1.9) |
| Smoking—n (%) |  |  |
| Past or current smoker | 50 (22.6) | 26 (25.2) |
| Never smoker | 171 (77.4) | 77 (74.8) |
| Travel within the past 3 months—n (%) |  |  |
| USA | 103 (46.6) | 55 (53.4) |
| Asia | 6 (2.7) | 3 (2.9) |
| Europe | 12 (5.4) | 9 (8.7) |
| Hospitalized—n (%) | 17 (7.7) | 8 (7.8) |
| Interval between symptom start and plasma donation (days)—mean (SD) |  | 45.1 (8.02) |
| Interval between symptom end and plasma donation (days)—mean (SD) |  | 33.3 (6.47) |
| ABO—n (%) |  |  |
| A positive |  | 35 (35.7) |

|  |  |
| --- | --- |
| A negative | 5 (5.1) |
| B positive | 13 (13.3) |
| B negative | 0 |
| AB positive | 4 (4.1) |
| AB negative | 0 |
| O positive | 35 (35.7) |
| O negative | 6 (6.1) |
| Anti-RBD antibody titer—mean (range, SD) | 522.5 (0-3892.3, 637.3) |
| Anti-Spike antibody titer—mean (range, SD) | 543.1 (0-3288.7, 564.8) |

BMI, body mass index; COPD, chronic obstructive pulmonary disease; OSA, obstructive sleep apnea; RBD, receptor binding domain. 103 plasma donors had donated plasma at date of manuscript submission with the remainder scheduled to donate in the upcoming weeks.

